# TRAUMATIC MICROHEMORHAGES ARE NOT SYNONYMOUS WITH AXONAL INJURY

**DOI:** 10.1101/2024.10.21.24315697

**Authors:** Rhonda Mittenzwei, Heather Maioli, Karinn Sytsma, Amanda Kirkland, C. Dirk Keene, Ramon Diaz-Arrastia, Christine Mac Donald, Chiara Maffei, Brian L. Edlow, Amber L. Nolan

**Author notes:** co-first authors (Rhonda Mittenzwei, Heather Maioli). co-last authors (Brian L. Edlow, Amber L. Nolan).

## Abstract

Diffuse axonal injury (DAI) is caused by acceleration-deceleration forces during trauma that shear white matter tracts. Susceptibility-weighted MRI (SWI) identifies microbleeds that are considered the radiologic hallmark of DAI and are used in clinical prognostication. However, this assumption is limited by a lack of systematic radiologic-pathologic correlation studies. Here, we performed *ex vivo* SWI on three brains from patients who died after severe TBI and assessed axonal injury around SWI microbleeds using immunohistochemistry to the amyloid beta precursor protein. Axonal injury was present in 28/44 microbleeds (64%), undermining the assumed pathophysiologic 1:1 link between traumatic vascular and axonal injury.

## INTRODUCTION

Traumatic microbleeds are a hallmark finding in patients with diffuse axonal injury (DAI).^1,2^ Rotational shearing forces that disrupt the cerebral microvasculature are likely to tear axons,^3^ such that the presence of a microbleed is typically interpreted as evidence of “hemorrhagic DAI.”^4^ Accordingly, in clinical practice, the presence of microbleeds factors heavily into prognostication, because of the assumption that axons, and hence neural networks,^5^ are disrupted.

Yet emerging evidence has called the pathophysiologic link between traumatic microbleeds and DAI into question. When comparing vascular injury, as detected by susceptibility-weighted imaging (SWI) or other magnetic resonance imaging (MRI) measures of vascular function, to the burden of DAI, as detected by diffusion MRI, several studies suggest that vascular injury and traumatic microbleeds may be dissociable from DAI.^6-8^ In addition, two correlative pathoradiologic studies of postmortem human tissue similarly suggest that axons may remain intact within and around traumatic microbleeds.^9,10^ However, systematic quantitative radiographic-pathologic correlation has not been performed to examine this relationship.

Elucidating the pathophysiological relationship between microbleeds and DAI is not only of neuroscientific interest, but also has immediate clinical relevance to patients with traumatic brain injury (TBI), for whom traumatic microbleeds are weighed heavily in prognostication. Indeed, the global burden of traumatic microbleeds,^9^ as well as the number of traumatic microbleeds in specific brain regions,^11^ such as the brainstem,^12^ have shown associations with long-term functional outcomes after TBI. Yet reports of functional recovery in patients with high numbers of global microbleeds,^13^ and multiple brainstem microbleeds,^14^ have reignited the debate about whether diffuse microbleeds are inextricably linked to a high degree of debilitating DAI.

To elucidate the relationship between traumatic microbleeds and DAI, we performed *ex vivo* SWI on three brains donated for research by patients who died less than two weeks after severe TBI. We used *ex vivo* SWI to guide histopathological sampling of white matter traumatic microbleeds and assessed axonal injury around these microbleeds using quantitative analysis of immunohistochemistry to the amyloid beta precursor protein (APP). Our observations generate new insights into the dissociation of traumatic microbleeds from DAI, weakening the pathophysiologic link between these entities and providing the basis for a reevaluation of prognostic models that rely upon traumatic microbleeds.

## METHODS

In this neuropathological case series, three brain donors with a history of severe TBI with a known mechanism of injury and death 1-2 weeks after injury were chosen for evaluation from the University of Washington Biorepository and Integrated Neuropathology laboratory. These cases (Table 1) included two from the Ex Vivo Connectomics study at Massachusetts General Hospital and one from the Brain Oxygen Optimization in Severe TBI Phase-3 (BOOST3)/Transforming Research and Clinical Knowledge in Traumatic Brain Injury (TRACK-TBI) study at the University of Pennsylvania. All studies were approved by the Massachusetts General Hospital, University of Pennsylvania, and University of Washington Institutional Review Boards and School of Medicine Compliance office. Informed consent for brain donation was provided by surrogate (next of kin) decision-makers.

**TABLE 1.**
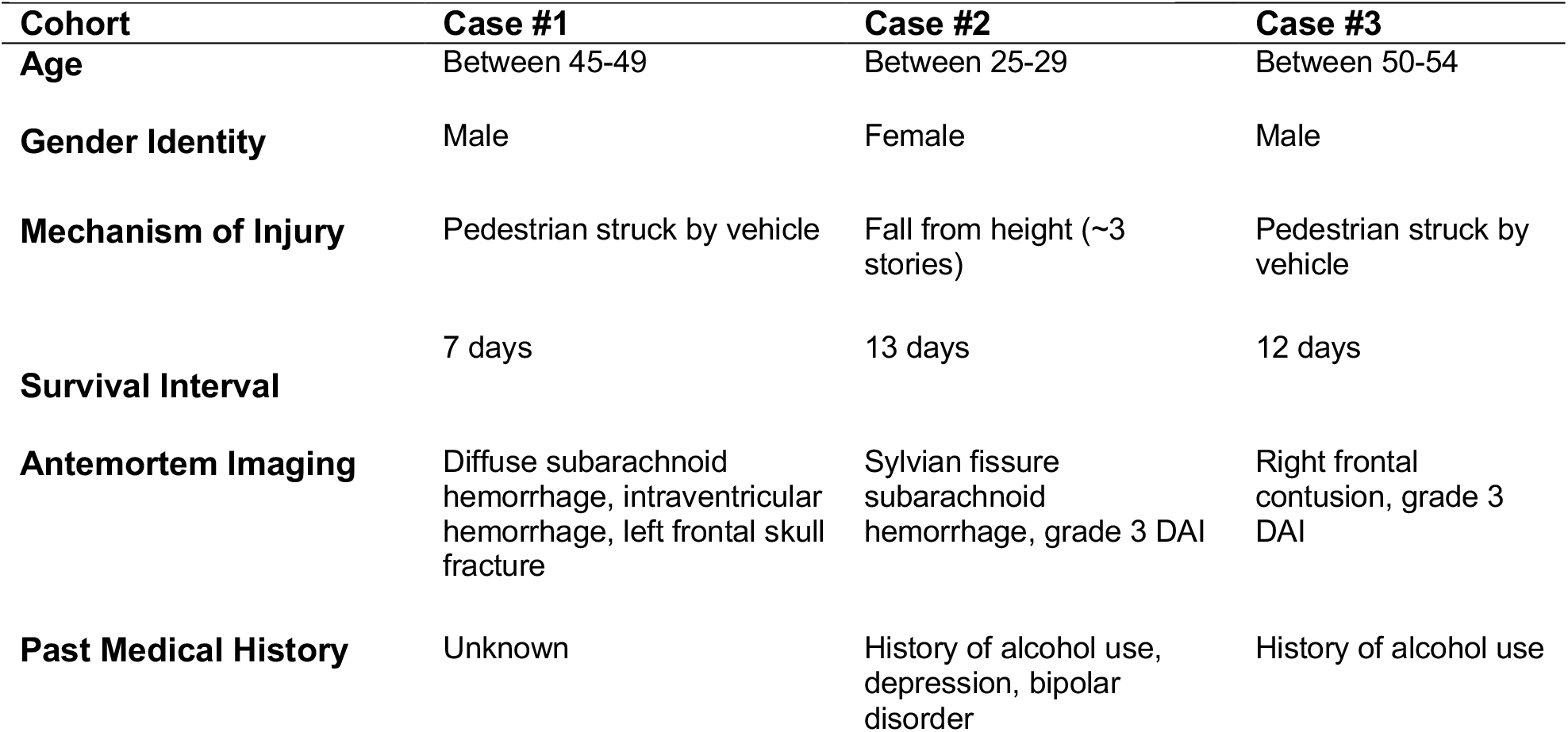
Clinical Features of Brain Donors. diffuse axonal injury (DAI)

| Cohort | Case #1 | Case #2 | Case #3 |
| --- | --- | --- | --- |
| Age | Between 45-49 | Between 25-29 | Between 50-54 |
| Gender Identity | Male | Female | Male |
| Mechanism of Injury | Pedestrian struck by vehicle | Fall from height (~3 stories) | Pedestrian struck by vehicle |
| Survival Interval | 7 days | 13 days | 12 days |
| Antemortem Imaging | Diffuse subarachnoid hemorrhage, intraventricular hemorrhage, left frontal skull fracture | Sylvian fissure subarachnoid hemorrhage, grade 3 DAI | Right frontal contusion, grade 3 DAI |
| Past Medical History | Unknown | History of alcohol use, depression, bipolar disorder | History of alcohol use |

Each donor underwent *ex vivo* MRI with the SWI sequence, followed by a comprehensive neuropathologic evaluation, as previously described.^15^ To guide pathological sampling of microbleeds, coronal brain slices of the cerebral hemispheres were spatially matched to coronal images from the *ex vivo* SWI dataset (Fig.1A-B). All regions with a white matter microbleed on SWI (defined as a punctate hypointense lesion at least 3 voxels in size located in at least 2 consecutive images at 500 μm resolution) were sampled (n = 34 blocks across the 3 cases) and then stained with hematoxylin and eosin (H&E) and APP on a Biocare autostainer (MAB348 Sigma-Aldrich, 1:75 dilution, epitope retrieval time 15 min) (Fig.1C-D).

**Fig. 1.**
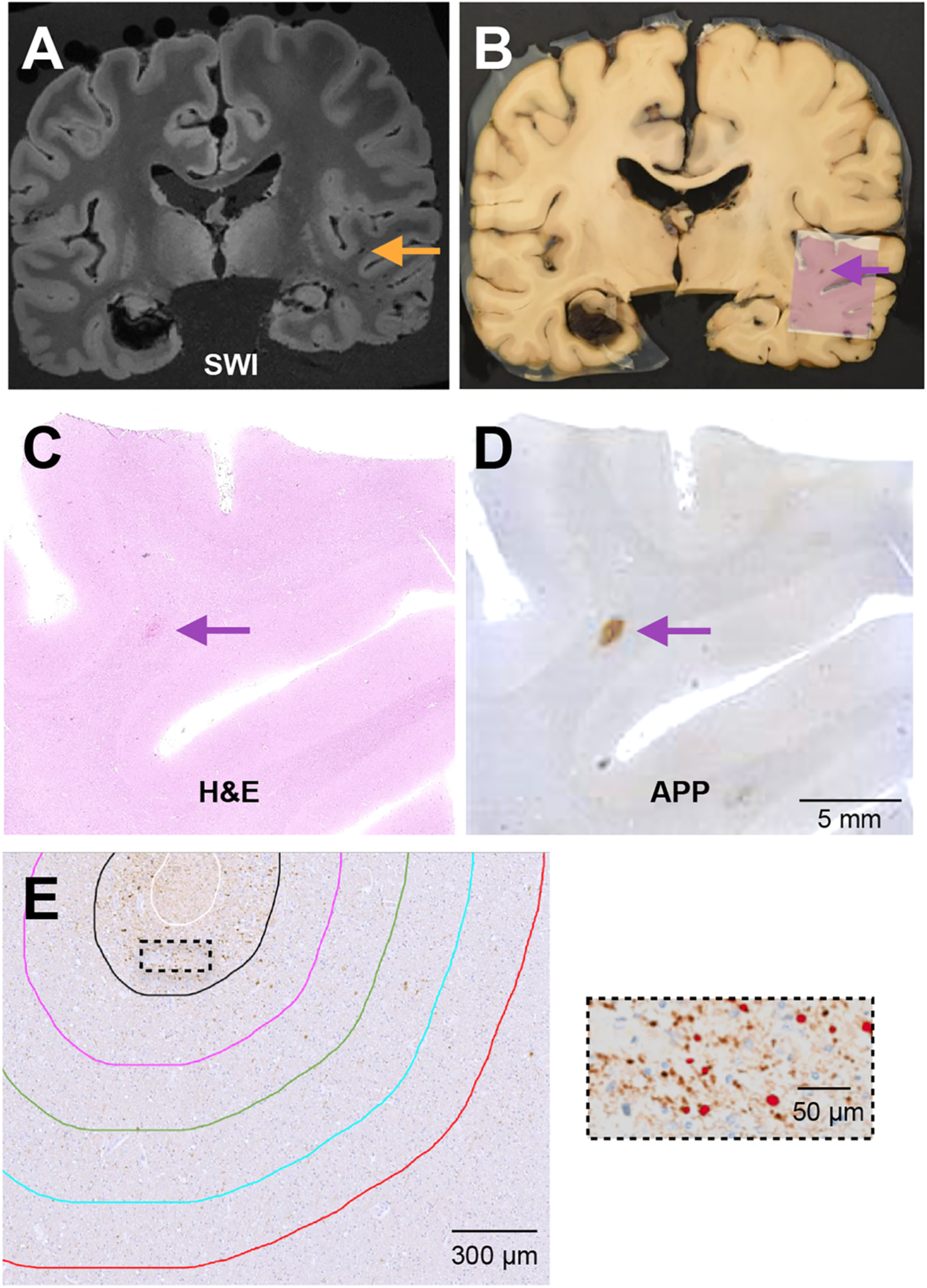
Methods to analyze axonal injury surrounding susceptibility weighted MR imaging (SWI)-identified traumatic microbleeds. A) Example coronal SWI image with traumatic microbleed denoted by orange arrow with corresponding gross brain slice (B). The same traumatic microbleed is denoted by a purple arrow and an image of the hematoxylin and eosin (H&E) slide is overlying the sampled region. C) The same H&E image from the sampled region and the sequential amyloid precursor protein immunohistochemical stained slide in D. E) Annotations for the regions of analysis performed in Halo. White line indicates the tracing around the microhemorrhage. Black, pink, green, blue and red lines represent 200-micron concentric rings up to 1 mm from the microbleed. The dashed box is shown at higher power on the right with the Halo algorithm to identify axonal swellings shown in red.

The presence of a microbleed was confirmed on H&E stain based on the presence of red blood cells outside the vascular wall and infiltrating into tissue. We manually traced each microbleed on the APP-stained slides using the HALO® platform (Indica Labs) (Fig.1E). Only single microbleeds, at least 2 mm away from another microbleed, were included for analysis. Axonal injury was assessed within 200 μm concentric rings up to 1 mm from the microbleed by determining the presence and density of APP+ axonal swellings. For within-case controls, the same concentric rings for each microbleed were placed randomly in the white matter on the same slide and examined.

Axonal swellings on the APP stain were defined as an object >4 μm in diameter using a pre-determined intensity threshold for every image (Fig.1E). These values correspond to the size of axonal swellings that have been reported.^16,17^ Axonal injury was defined by the presence of >2 swellings within one ring; this threshold was chosen to reduce the false positive rate associated with the algorithm, rarely identifying inappropriate background staining. All microbleeds were also verified by a neuropathologist to confirm true APP+ axonal swelling. Microbleeds were excluded from analysis if too much background staining was present and interfered with the Halo imaging algorithm, as visually inspected by a neuropathologist. To determine whether the relationship between microbleeds and axonal injury depends on neuroanatomic location, we classified each microbleed by lobe, location within the white matter (i.e., superficial or deep), and proximity to contusion. The presence of axonal spheroids on H&E stain was also determined for every microbleed by a neuropathologist, blinded to the results of the APP analysis.

Statistical analyses were performed using GraphPad Prism 9 software. The relationship of axonal swellings as a function of distance was analyzed with a Friedman test. A Fischer’s exact test determined the association of APP+ axonal swellings with the presence of H&E axonal spheroids as well as neuroanatomic location and control sampling.

Unbiased clustering analysis of the density of APP+ axonal swellings was also performed to provide a more objective measure to classify microbleeds. A K-means clustering algorithm, which optimizes the sum of the square Euclidean distance between points, was performed in MATLAB (version R2022a) software on the density of APP+ swellings within the first 200-micron ring.

## RESULTS

Axonal injury was associated with 28/44 (64%) microbleeds, as defined by the presence of APP+ axonal swellings (Fig.2A). This proportion was substantially higher than that observed in the random analysis of white matter (5/44 randomly sampled regions, p<0.001, Fisher’s exact test). The number of microbleeds and percentage with axonal injury was variable across cases (Fig.2A). The density of APP+ axonal swellings decreased as a function of distance for microbleeds positive for axonal injury (Fig.2B), and the presence of axonal spheroids on H&E was strongly correlated with the presence of axonal injury detected by APP stain (Fig.2C). The neuroanatomic classification of microbleeds did not affect the relationship between microbleeds and axonal injury (Fig.2D).

**Fig. 2.**
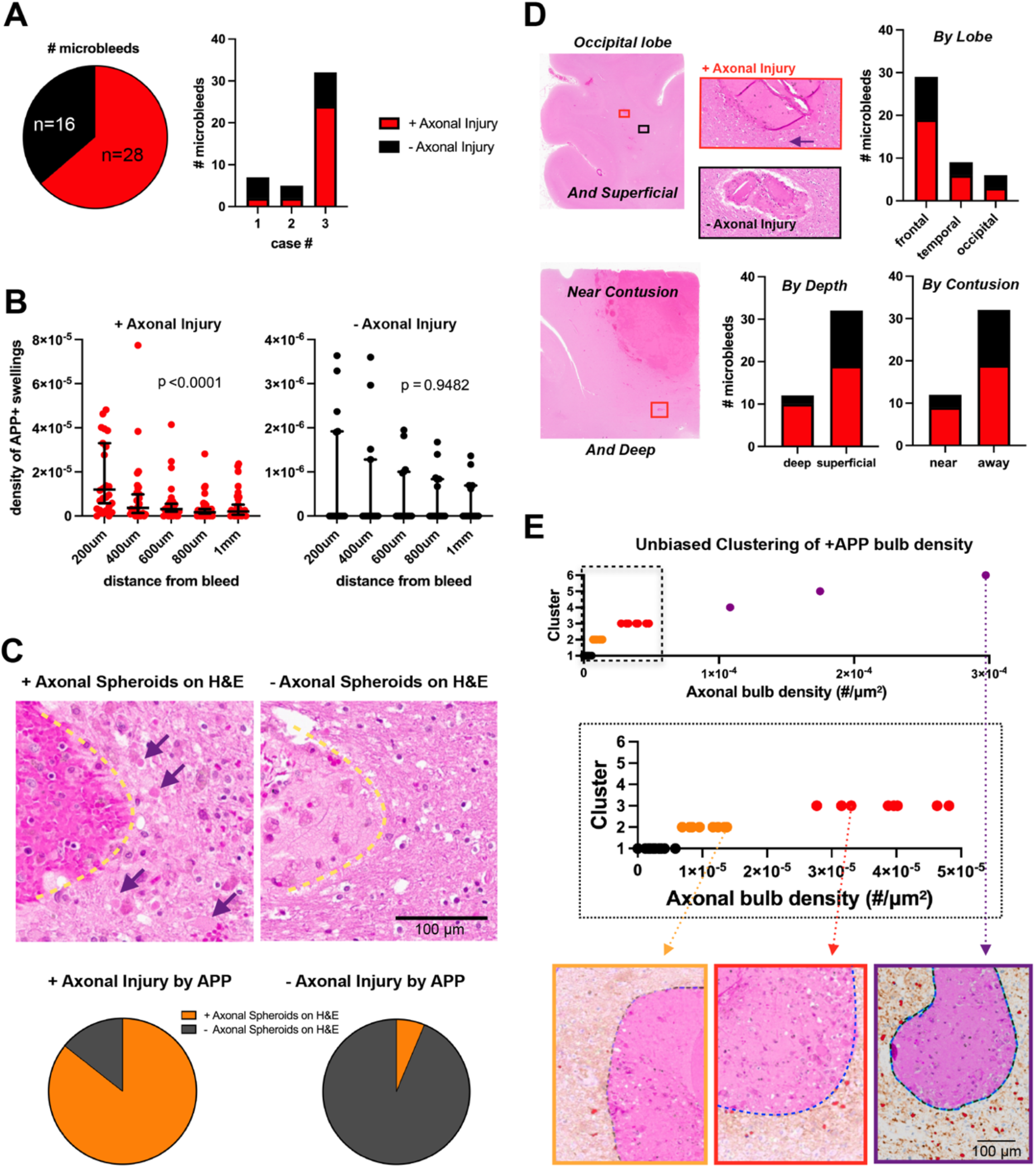
Axonal injury is not directly synonymous with traumatic microbleeds. A) The number of microbleeds with axonal injury identified with amyloid precursor protein (APP) (28/44) and the microbleed distribution across cases. B) The relationship between distance and APP+ axonal swellings in those considered positive for axonal injury on left (p < 0.0001, Friedman test) and without axonal injury on right (p = 0.9482, Friedman test). C) Representative traumatic microbleeds with axonal spheroids (purple arrows) on hematoxylin and eosin (H&E) stain on the left and without axonal spheroids on the right. 24/28 traumatic microbleeds positive for axonal injury on APP had axonal spheroids seen on H&E. While only 1/15 traumatic microbleeds without axonal injury on APP stain exhibited an axonal spheroid on H&E. Fischer’s exact test shows significant correlation (p < 0.0001) between axonal injury on APP stain and axonal spheroids on H&E. D) Representative images of microbleeds in the occipital lobe and near a contusion. The red boxes indicate axonal damage on APP stain and H&E, the black box indicates no axonal injury.. No significant relationship seen by location in lobe (p = 0.8045, Fischer’s exact test), superficial versus deep (p=0.1712, Fischer’s exact test), or proximity to contusion (p=0.4866, Fischer’s exact test). E) Unbiased clustering of the APP+ axonal swellings (bulbs) with the kmeans algorithm. There were three outliers with a very high density of APP+ swellings (purple), and then three other clusters with a minimal (black), mild (orange) and moderate (red) degree of injury.

Unbiased clustering analysis of the density of APP+ axonal swellings revealed three outliers with a very high density of APP+ swellings (purple, Fig.2E), and three other clusters with a minimal (black), mild (orange) and moderate (red) degree of injury (Fig.2E). When this unbiased tool is used to define axonal injury, the minimal group that might be inferred to represent microbleeds without significant axonal injury includes 25 microbleeds (57%), raising even higher the number of microbleeds “negative” for axonal injury.

## DISCUSSION

We performed systematic quantitative radiologic-pathologic analysis to determine the relationship of axonal injury to traumatic microbleeds identified on SWI in three brain specimens donated by patients with acute severe TBI. We found that only a subset (64%) of microbleeds exhibit adjacent axonal injury. Our data also reveal a variable severity of axonal injury associated with microbleeds, as the majority of microbleeds with adjacent axonal injury have very few APP+ axonal swellings.

While this is the first systematic quantitative analysis of traumatic microbleeds and axonal injury, our data aligns with prior observations. In a patient with severe TBI who died 7 months after injury, microbleeds on post-mortem MRI were found to exhibit iron deposition and hemosiderin-laden macrophages without axonal injury, as indicated by a lack of axonal spheroids, beading or APP positivity.^9^ In a patient with traumatic coma who died 3 days after injury, post-mortem MRI and pathological anlaysis revealed a cingulum bundle hemorrhage without associated axonal injury.^10^ In a swine model of head rotational acceleration, inverstigators observed fibrinogen extravasation, a marker of blood-brain barrier disruption, in the absence of axonal injury.^18^ Fibrinogen extravasation only co-occurred in ∼60% of the area of axonal injury,^18^ consistent with our observations in humans that vascular and axonal injury are not inextricably linked.

While our results do not support a direct 1:1 relationship with microbleed pathology and axonal injury, microbleed pathology alone may still be clinically relevant for some patients. Their presence on neuroimaging was an independent predictor of disability in a cohort of largely mild TBI cases^9^; in contrast, in a moderate-severe TBI cohort only diffusion tensor imaging measures of axonal injury correlated with poorer cognitive and functional outcomes while no difference in cognitive performance was found in comparing patients with and without microbleeds^8^. Axonal injury itself is also not necessarily permanent or incapable of recovery; up to ∼10% of axonal swellings thought to be terminal transections were revealed with 3D histological analysis to be intact and potentially salvageable.^19^ Our findings, in the context of these prior studies, thus highlight the need to develop clinical tools that detect preserved axons and more accurately predict recovery in patients with TBI^20,21^.

Why some microbleeds are associated with axonal injury and others are not is unclear. We examined the location of microbleeds by superficial versus deep white matter, lobe and whether adjacent to contusion, and none of these metrics were significantly associated with the presence of axonal injury around microbleeds. An imaging study suggested that microbleeds in the midsagittal region (cingular cortex, parasagittal white matter, and corpus callosum) are more likely to be associated with axonal injury,^6^ but most microbleeds in this study were in lobar regions of the brain, not the subcortical regions that are more relevant to prognostication in severe TBI.^11,12^ The specific white matter pathways that a vessel is associated with might also influence the likelihood of axonal injury, with long-ranging pathways being perhaps more vulnerable. This hypothesis will require testing with *ex vivo* diffusion MRI tractography, as the white matter pathway that an axon/vessel is involved with cannot be determined with pathology alone. Finally, cellular responses to the microbleed could variably affect axonal integrity due to the heterogenous populations of astrocytes and microglia that have been found across grey and white matter regions of the brain.^22-24^

Our study has several limitations. The three patients with severe TBI died within 1-2 weeks after injury, raising the possibility that some microbleeds and/or associated axonal injury may not be attributable to the trauma itself, but might be secondary to other processes such as ischemia and swelling that occur after severe TBI. We also only analyzed “single” microbleeds that were > 2 microns away from other microbleeds on microscopy. Finally, the *ex vivo* SWI and subsequent sampling did not include the brainstem, which is routinely removed before imaging in our protocol. Future studies will need to incorporate more acute cases, a greater number of regions including the brainstem, and clusters of microbleeds to determine if the principles observed here are generalizable.

In summary, our systematic quantitative evaluation of microbleeds weakens the dogma of “hemorrhagic diffuse axonal injury”, indicating that microbleeds and axonal injury are not as spatially correlated as previously thought. These results provide the basis for a reevaluation of prognostic models in TBI that strongly rely upon traumatic microbleeds identified on imaging.

## Data Availability

All data produced in the present study are available upon reasonable request to the authors

## ACKNOWLEDGEMENTS

This work was supported by NIH/NINDS (K08NS114170, R01NS138257, R21NS109627, R01NS091618, U01NS086090, U54NS115322, U01NS137500, U24NS135561, RF1NS115268, U01NS086625, U01NS137484), NIH/NIA (P30AG066509, AG066509, U24AG072458,), NIH Director’s Office (DP2HD101400), the United States Department of Defense (W81XWH-21-S-TBIPH2), the VA Advanced Fellowship in Mental Illness Research and Treatment, the Chen Institute MGH Research Scholar Award, and the Nancy and Buster Alvord Endowment. We thank the donors and their families for their contribution to science.

## DECLARATION OF INTERESTS

The authors declare no competing interests.

## AUTHOR CONTRIBUTIONS

C.D.K., R.D.A., and B.L.E. were involved in the brain donation of these cases. R.M, H.M., C.M. and C.M.D. identified the microhemorrhages on imaging and sampled the brain tissue. A.K. performed the histology and staining. R.M., H.M., K.S., and A.L.N. performed the Halo annotations and analysis. B.L.E., R.D.A., C.M.D., C.M., and A.L.N. contributed to the design and approach of the study. A.L.N and B.L.E. wrote the paper.

## Notes

### Competing Interest Statement

The authors have declared no competing interest.

### Author Declarations

All studies were approved by the Massachusetts General Hospital, University of Pennsylvania, and University of Washington Institutional Review Boards and School of Medicine Compliance office. Informed consent for brain donation was provided by surrogate (next of kin) decision-makers.

## REFERENCES

1. Courville CB. Traumatic intracerebral hemorrhages, with special reference to the mechanics of their production. Bull Los Angel Neuro Soc 1962;27:22–38. (https://www.ncbi.nlm.nih.gov/pubmed/13881792).

2. Povlishock JT, Christman CW. The pathobiology of traumatically induced axonal injury in animals and humans: a review of current thoughts. J Neurotrauma 1995;12(4):555–64. DOI: 10.1089/neu.1995.12.555.

3. Bailey P. Traumatic apoplexy. Med Rec 1904;66:528 – 529..

4. Tong KA, Ashwal S, Holshouser BA, et al. Diffuse axonal injury in children: clinical correlation with hemorrhagic lesions. Ann Neurol 2004;56(1):36–50. DOI: 10.1002/ana.20123.

5. Sharp DJ, Scott G, Leech R. Network dysfunction after traumatic brain injury. Nat Rev Neurol 2014;10(3):156–66. DOI: 10.1038/nrneurol.2014.15.

6. Andreasen SH, Andersen KW, Conde V, et al. Limited Colocalization of Microbleeds and Microstructural Changes after Severe Traumatic Brain Injury. J Neurotrauma 2020;37(4):581–592. DOI: 10.1089/neu.2019.6608.

7. Haber M, Amyot F, Lynch CE, et al. Imaging biomarkers of vascular and axonal injury are spatially distinct in chronic traumatic brain injury. J Cereb Blood Flow Metab 2021;41(8):1924–1938. DOI: 10.1177/0271678X20985156.

8. Jolly AE, Balaet M, Azor A, et al. Detecting axonal injury in individual patients after traumatic brain injury. Brain 2021;144(1):92–113. DOI: 10.1093/brain/awaa372.

9. Griffin AD, Turtzo LC, Parikh GY, et al. Traumatic microbleeds suggest vascular injury and predict disability in traumatic brain injury. Brain 2019;142(11):3550–3564. DOI: 10.1093/brain/awz290.

10. Edlow BL, Haynes RL, Takahashi E, et al. Disconnection of the ascending arousal system in traumatic coma. J Neuropathol Exp Neurol 2013;72(6):505–23. DOI: 10.1097/NEN.0b013e3182945bf6.

11. Bianciardi M, Izzy S, Rosen BR, Wald LL, Edlow BL. Location of Subcortical Microbleeds and Recovery of Consciousness After Severe Traumatic Brain Injury. Neurology 2021;97(2):e113–e123. DOI: 10.1212/WNL.0000000000012192.

12. Izzy S, Mazwi NL, Martinez S, et al. Revisiting Grade 3 Diffuse Axonal Injury: Not All Brainstem Microbleeds are Prognostically Equal. Neurocrit Care 2017;27(2):199–207. DOI: 10.1007/s12028-017-0399-2.

13. Edlow BL, Giacino JT, Hirschberg RE, Gerrard J, Wu O, Hochberg LR. Unexpected recovery of function after severe traumatic brain injury: the limits of early neuroimaging-based outcome prediction. Neurocrit Care 2013;19(3):364–75. DOI: 10.1007/s12028-013-9870-x.

14. Young MJ, Sanders WR, Marujo R, Bodien YG, Edlow BL. Return to Work Within Four Months of Grade 3 Diffuse Axonal Injury. Neurohospitalist 2022;12(2):280–284. DOI: 10.1177/19418744211051459.

15. Latimer CS, Melief EJ, Ariza-Torres J, et al. Protocol for the Systematic Fixation, Circuit-Based Sampling, and Qualitative and Quantitative Neuropathological Analysis of Human Brain Tissue. Methods Mol Biol 2023;2561:3–30. DOI: 10.1007/978-1-0716-2655-9_1.

16. Nolan AL, Petersen C, Iacono D, et al. Tractography-Pathology Correlations in Traumatic Brain Injury: A TRACK-TBI Study. J Neurotrauma 2021;38(12):1620–1631. DOI: 10.1089/neu.2020.7373.

17. Johnson VE, Stewart W, Weber MT, Cullen DK, Siman R, Smith DH. SNTF immunostaining reveals previously undetected axonal pathology in traumatic brain injury. Acta Neuropathol 2016;131(1):115–35. DOI: 10.1007/s00401-015-1506-0.

18. Johnson VE, Weber MT, Xiao R, et al. Mechanical disruption of the blood-brain barrier following experimental concussion. Acta Neuropathol 2018;135(5):711–726. DOI: 10.1007/s00401-018-1824-0.

19. Weber MT, Arena JD, Xiao R, Wolf JA, Johnson VE. CLARITY reveals a more protracted temporal course of axon swelling and disconnection than previously described following traumatic brain injury. Brain Pathol 2019;29(3):437–450. DOI: 10.1111/bpa.12677.

20. Sanders WR, Barber JK, Temkin NR, et al. Recovery Potential in Patients Who Died After Withdrawal of Life-Sustaining Treatment: A TRACK-TBI Propensity Score Analysis. J Neurotrauma 2024. DOI: 10.1089/neu.2024.0014.

21. Bodien YG, Allanson J, Cardone P, et al. Cognitive Motor Dissociation in Disorders of Consciousness. N Engl J Med 2024;391(7):598–608. DOI: 10.1056/NEJMoa2400645.

22. Amor S, McNamara NB, Gerrits E, et al. White matter microglia heterogeneity in the CNS. Acta Neuropathol 2022;143(2):125–141. DOI: 10.1007/s00401-021-02389-x.

23. Tan YL, Yuan Y, Tian L. Microglial regional heterogeneity and its role in the brain. Mol Psychiatry 2020;25(2):351–367. DOI: 10.1038/s41380-019-0609-8.

24. Mazumder AG, Jule AM, Cullen PF, Sun D. Astrocyte heterogeneity within white matter tracts and a unique subpopulation of optic nerve head astrocytes. iScience 2022;25(12):105568. DOI: 10.1016/j.isci.2022.105568.

